# Vomiting during body surface gastric mapping testing

**DOI:** 10.1101/2025.10.08.25337635

**Authors:** Sam Simmonds, Daphne Foong, Gabriel Schamberg, Gen Johnston, Vincent Ho, Anthony Hobson, Armen A. Gharibans, Christopher N. Andrews, Greg O’Grady, Stefan Calder

## Abstract

**Background:** Vomiting is a symptom of various gastrointestinal (GI) disorders and may invalidate gastric emptying tests. Body surface gastric mapping (BSGM) is a clinical test to assess motor *vs*. sensory contributors to GI symptoms.

**Aims:** While previous studies have observed myoelectrical dysrhythmias associated with vomiting, the effects of vomiting on BSGM testing have not been defined.

**Methods:** A large clinical database of de-identified BSGM tests was queried for vomiting events, noted by symptom markers via an integrated symptom reporting App. Tests with pre-meal or >2 vomiting events were excluded. Spectrograms and clinical reports were qualitatively assessed. Key BSGM metrics, including the Gastric Alimetry Rhythm Index (GA-RI) and BMI-adjusted amplitude, were interrogated in 5 min epochs for quantitative analysis.

**Results:** A total of 49 vomit events were included. Vomiting typically had little effect, though was sometimes characterised by small, temporary decreases in BMI-adjusted amplitude or GA-RI. Prolonged periods (> 10 mins) of low amplitude were observed in 4 cases (8%). A mixed effects model revealed a transient decrease in GA-RI in the 5 mins before (Δ = −0.27; *p* < 0.001) and after vomiting (Δ = −0.21; *p* = 0.014), but not in subsequent periods (all *p* > 0.05). Other metrics were unaffected. Nausea, bloating, and excessive fullness symptoms decreased following vomiting (all *p* < 0.05).

**Conclusions:** Transient amplitude and rhythm decreases were observed concurrent to vomiting, but subsequently normalised. While additional considerations may be required during test interpretation, the overall impact of vomiting on BSGM test interpretation is minimal.

## Introduction

Nausea and vomiting may be debilitating symptoms in many chronic gastroduodenal symptoms, and are cardinal features of gastroparesis,(Cherian and Parkman 2012) cannabis hyperemesis syndrome,(Hasler et al. 2022) and chronic nausea and vomiting syndromes.(Lacy et al. 2018) Nausea and vomiting mechanisms are multifactorial, and may be induced by central(Miller 1999) and peripheral(Babic and Browning 2014) mechanisms.

Gastric emptying testing is widely performed to investigate chronic nausea and vomiting symptomatology. However, vomiting episodes during gastric emptying tests will typically invalidate the results.(Farrell 2019) Gastric emptying tests may therefore be ill-suited to be the mainstay diagnostic test for patients with frequent and predominant vomiting symptoms, compromising healthcare efficiency.

Body surface gastric mapping (BSGM) via Gastric Alimetry® (Auckland, New Zealand) is a non-invasive tool for measuring gastric myoelectrical activity alongside validated real-time symptom logging.(Foong et al. 2023) BSGM is used to classify patients with abnormal meal responses and underlying gastric myoelectrical dysfunction, to direct care towards specific motor or sensory pathologies.(Gharibans et al. 2022; Wang et al. 2024; Varghese et al. 2025a) However, the effects of vomiting on BSGM data have not yet been characterised. Furthermore, it is important to assess whether vomiting may compromise the interpretability of Gastric Alimetry results.

The Gastric Alimetry system measures gastric slow-wave activity, together with contractility, from the body surface using a dense array of 66 electrodes (8 ⍰ 8, +2 reference; inter-electrode spacing 20 mm).(Gharibans et al. 2023) BSGM recording across pre- and post-prandial periods enables insight into gastric function in the fasted and fed states, as well as characterising the meal response. BSGM generates summative metrics, including the Gastric Alimetry Rhythm Index (GA-RI; range 0-1), principal gastric frequency (PGF; cycles per minute [cpm]), and BMI-adjusted amplitude (µV), to characterise the gastric myoelectrical activity.(O’Grady et al. 2023a) Interpretation of these metrics against established normative reference intervals can be used to define underlying gastric electrophysiology and digestive function.(Foong et al. 2023) For example, the presence of spectral scattering (GA-RI < 0.25) indicates underlying dysrhythmia that is considered to be associated with depletion of interstitial cells of Cajal, a common pathological hallmark in nausea and vomiting syndromes.(O’Grady et al. 2012; Angeli et al. 2015)

The aim of this study was therefore to characterise the myoelectrical activity before and after a logged vomiting event on BSGM, and assess the impact of vomiting on overall BSGM interpretation.

## Methods

### Study Procedure and Inclusion Criteria

An existing database of over 2595 de-identified BSGM cases were queried for Gastric Alimetry tests that included vomiting episodes. All patients provided informed consent for data collection and anonymized data use for research purposes at the time of testing, with cases pooled from three regions: Auckland (New Zealand), Calgary (Canada), and Western Sydney (Australia); ethical approvals: AH1130, REB19-1925, and H13541.

This study specifically aimed to characterize the local temporal myoelectrical changes occurring around a logged vomiting event and to assess subsequent trends in gastric activity. To ensure an adequate period for observing this recovery and to clearly delineate the local effects without confounding influences from multiple closely spaced emetic events, the analysis was restricted to tests with 1 or 2 recorded vomiting episodes.

Patients aged 18 years or older were included for analysis if they had standard test protocols: 30 min baseline recording, followed by a standardised meal consisting of Ensure [232 kcal, 250 mL; Abbott Nutrition, USA] and an oatmeal energy bar or similar [250 kcal, 5 g fat, 45 g carbohydrate, 10 g protein, 7 g fibre; Clif Bar & Company, USA], and a 4 hr postprandial recording. Recordings with significant artefacts (> 70 %; independent of vomiting), missing significant data (> 1 hr), non-standard meals (*e*.*g*., scintigraphy or liquid meals), pre-prandial vomiting, or greater than 2 vomit markers (in order to avoid inclusion of rumination syndrome) were not included for analysis. All tests that met these inclusion criteria were included for qualitative analysis.

### Quantitative and Qualitative Analysis

Gastric Alimetry reports were screened for vomit-related motion artefacts and general test quality, as previously detailed.(Foong et al. 2023) Gastric Alimetry phenotypes were determined based on normative intervals for spectral metrics, as previously described.(Gharibans et al. 2022; O’Grady et al. 2023b; O’Grady et al. 2023a)

BSGM metrics were analysed in six 5 min epochs (three prior to and three following the logged vomiting event) to investigate the local effects of vomiting on BSGM metrics and the presence of vomiting-induced motion artefacts. Vomit events were excluded from quantitative analysis if they occurred before meal consumption or within 15 min of the conclusion of meal consumption, to ensure that analysed epochs did not contain fasting data, which tends to have large variability likely due to the presence of migrating motor complexes.(Deloose et al. 2012) Additionally, BSGM metrics were calculated with the exclusion of the 5 min before and after the vomiting event and compared to metrics derived from the full dataset, to evaluate the overall impact of a vomiting event on BSGM metrics.

Symptoms were captured throughout the BSGM recording using the Gastric Alimetry symptom logging App, as validated by Sebaratnam *et al*.*(Sebaratnam et al. 2022; Sebaratnam et al. 2023)* Symptoms included nausea, bloating, excessive fullness, heartburn, stomach burn, and upper gut pain.

### Statistical Analysis

Linear mixed effects models were used to assess changes in symptoms, BMI-adjusted amplitude, and GA-RI across epochs surrounding the vomiting event. Each model included a random intercept for participants to account for repeated measures. Epochs were treated as a categorical fixed effect, with the earliest time point (15 min prior to logged vomiting event) used as the reference. PGF was excluded from analysis within 5 min epochs, as its calculation is not robust to this temporal resolution. Results are reported as estimated changes from the reference epoch (Δ), corresponding to the model’s fixed-effects estimates, along with 95 % confidence intervals (CIs) and *p*-values. Mann-Whitney U tests were used to confirm statistical differences. Results are reported as mean ± SD unless otherwise stated, and a significance threshold of 0.05 was used for all tests. All analyses were performed using Python 3.9.

## Results

172 (7%) tests containing at least one vomiting event were returned from the database query. A total of 49 logged vomiting events (from 42 individuals) were included for qualitative analysis, after applying the exclusion criteria (median age 30 years [range 19 – 79], BMI 24.5 kg/m^2^ ± 6.0 kg/m^2^ [mean ± SD], 73 % female). These individuals had a median meal consumption of 80 % (CI: 66 % – 83 %).

Patients included for qualitative analysis had a variety of Alimetry spectral phenotypes: 32 (76%) were spectrally normal, 5 (12%) were dysrhythmic, 4 (10%) had a BMI-adjusted amplitude outside the reference intervals (1 (2%) high, 3 (7%) low), and 1 (2%) had low frequency. After applying the quantitative analysis exclusion criteria, 32 logged vomiting events from 32 individuals were included. Individuals included for quantitative analysis had mean BMI-adjusted amplitude 40.1 µV ± 22.5 µV, PGF 3.00 cpm ± 0.21 cpm, and GA-RI 0.44 ± 0.22.

### Effects on Clinical Interpretation

We investigated the complete effect of vomiting on BSGM by analysing both the immediate “vomiting signature” (± 5 min period surrounding a logged event) and by quantitatively analysing the subsequent spectral analysis.

The majority of clinical reports (60 %) did not show a consistent BSGM signature surrounding logged vomiting events. In some reports, however, vomiting was associated with transient (*i*.*e*., < 10 min), localised alterations in the BSGM recording in several ways. Some patients experienced a drop in BMI-adjusted amplitude (12 %; **Fig. 1, 2C**), gastric rhythm stability (quantified as GA-RI) (20 %; **Fig. 2D**), or quiescence, where there was no distinct gastric slow wave signal (8 %; **Fig. 2A**). Additionally, artefacts associated with motion during vomiting (*i*.*e*., those not corrected by the automated noise cancellation) were observed in 3 cases (6 %; **Fig. 2B**).

**Fig. 1.**
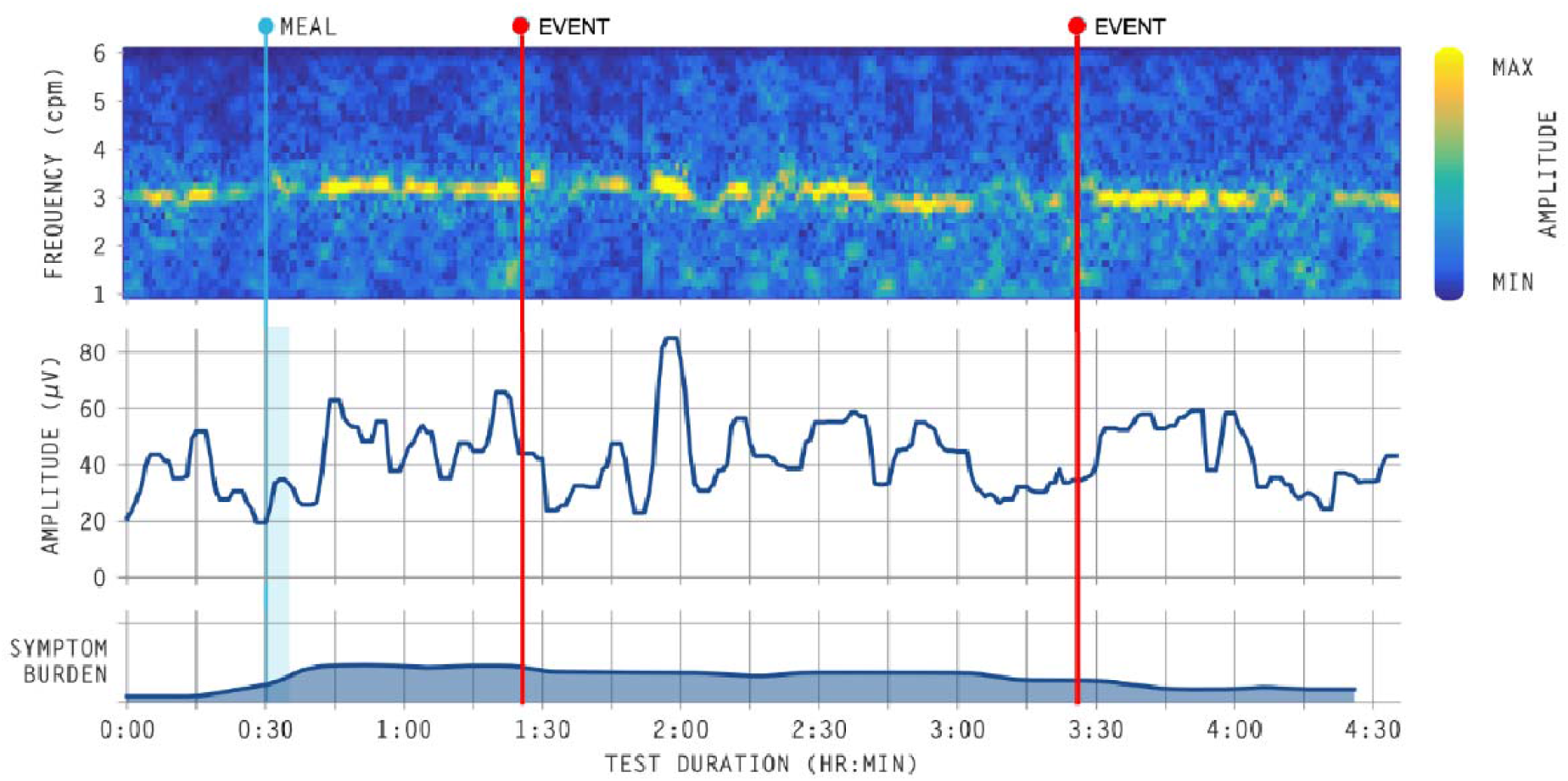
In most cases, vomiting did not significantly impact the quality of BSGM reports. In this example, BMI-adjusted amplitude decreases surrounding the vomiting events, but returns to normative values within ~30 min. Event markers denote when vomiting occurs

**Fig. 2.**
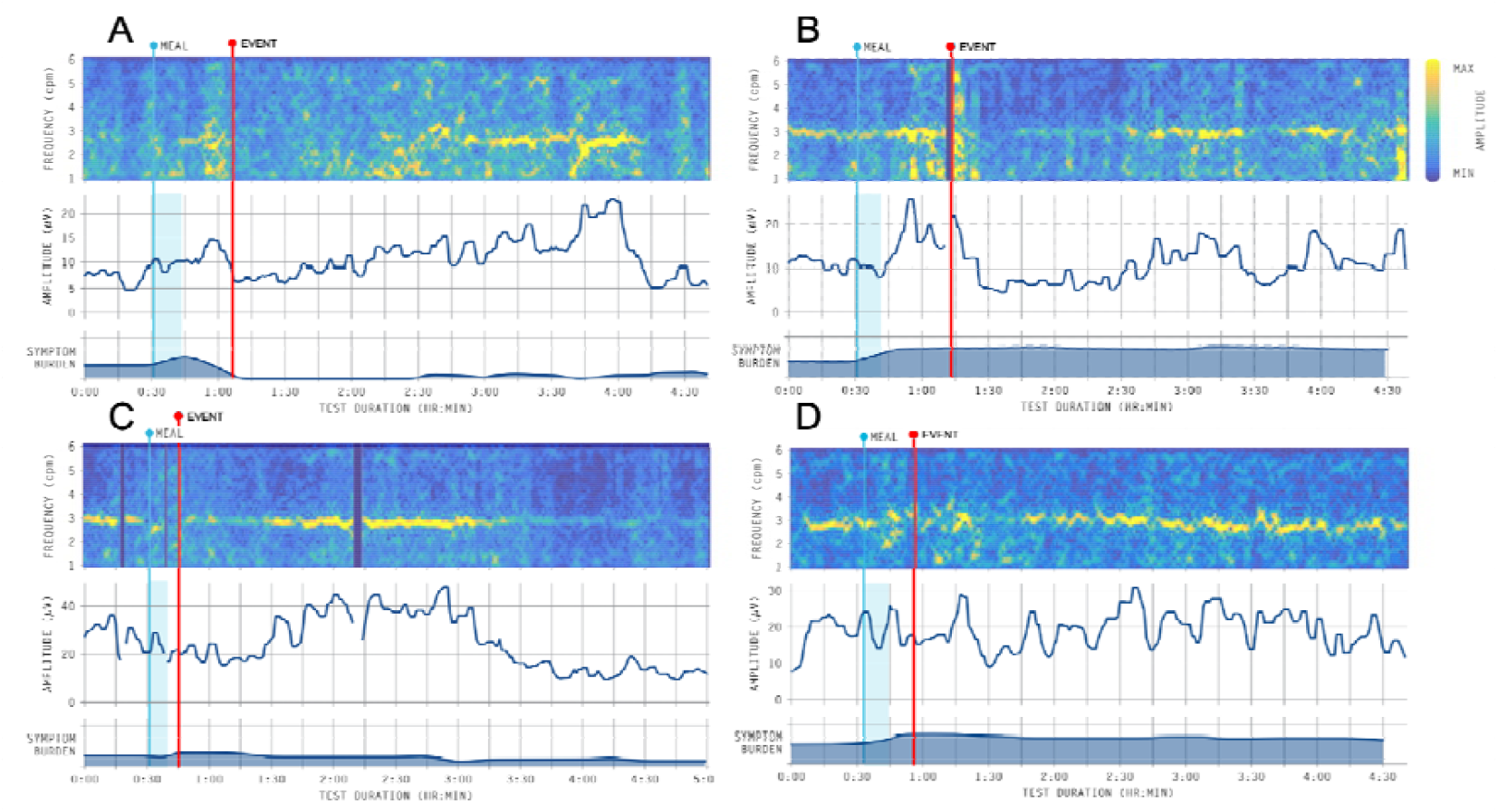
Vomiting has different presentations on BSGM results. *Prolonged effects:* a) Vomiting preceded a period of quiescence, which then transitioned into activity with low rhythm stability for ~30 min before returning to pre-vomit behaviour. b) Vomiting was associated with reduced amplitude and a delayed meal response, before returning to normal behaviour ~45 min after the vomiting event. c) Vomiting immediately after meal consumption may have delayed the meal response (increase in BMI-adjusted amplitude) by ~45 min. *Immediate effects:* Surrounding the vomiting event, vomiting was associated with transient b) noise, c) a drop in BMI-adjusted amplitude amplitude, or d) a reduction in gastric rhythm stability (visualised as a scattering on the frequency spectra). Event markers denote when vomiting occurs

Subsequent effects on the spectral pattern could theoretically reflect reduced stomach content volume and modified digestive function. In most cases, however, vomiting was not associated with apparent prolonged effects or changes in spectral morphology, such that clinical report interpretation was concluded to be functionally equivalent to cases without vomiting. An example case is presented in **Fig. 1**, which was classified with a normal spectral pattern despite BMI-adjusted amplitude deviations coinciding with the two logged vomiting episodes.

There were only four instances (8%) where visual inspection of the clinical reports suggested that vomiting may have altered the subsequent (*i*.*e*., persisting >10 min) bioelectrical behaviour captured by BSGM (**Fig. 2**). In each case, vomiting appeared to precede a period of low amplitude or relative quiescence. In one such case vomiting occurred ~30 min after meal consumption, where it was associated with a transient period of low rhythm stability (GA-RI < 0.25), before preceding a ~1 hr period of electrical quiescence, which finally resolved with the return of normal (pre-vomit) activity with stable rhythm stability (**Fig. 2B**). In another individual, the period of relative quiescence seemed to delay the onset of the meal response after vomiting (**Fig. 2A**). In all cases, BSGM activity returned to pre-vomit behaviour in ~1.5 hr or earlier, regardless of myoelectrical presentation before the vomiting (**Fig. 2A–C**). This can perhaps best be visualised with **Figs. 2A** and **2C**; the former expressing clear dysrhythmic behaviour following consumption of the meal and resolution of the vomiting-associated quiescence, while the latter demonstrates similar behaviour for a spectrally-normal stomach.

Most cases, however, were similar to **Fig. 1**, where pre- and post-vomiting behaviour were indistinguishable other than the transient effects immediately surrounding the logged event. This is further supported by the removal of epochs surrounding the logged vomiting event (± 5 min), which did not affect the overall post-prandial BMI-adjusted amplitude (38.81 µV ± 21.23 µV vs. 37.88 µV ± 20.77 µV; *p* = 0.726), PGF (3.03 cpm ± 0.24 cpm vs. 2.97 cpm ± 0.25 cpm; *p* = 0.125), or GA-RI (0.44 ± 0.19 vs. 0.43 ± 0.18; *p* = 0.806), indicating that these clinically-reported post-prandial BSGM metrics were not affected by vomiting episodes.

### Localised Effects

A mixed effects model of pre- and post-vomit epochs revealed several statistically significant changes that physiologically defined the vomiting period (**Table 1**). Compared to the reference period (15 min to 10 min prior to vomiting), there was a statistically significant decrease in GA-RI over the 5 min immediately before vomiting ( = −0.27; 95 % CI −0.41 to −0.13; *p* < 0.001), and in the epoch immediate after vomiting ( = −0.21; 95 % CI −0.38 to −0.04; *p* = 0.014), but not in subsequent periods (all *p* > 0.05), indicating that GA-RI changes were transient. There were no meaningful changes in BMI-adjusted amplitude across epochs (all *p* > 0.05).

**Table 1.**
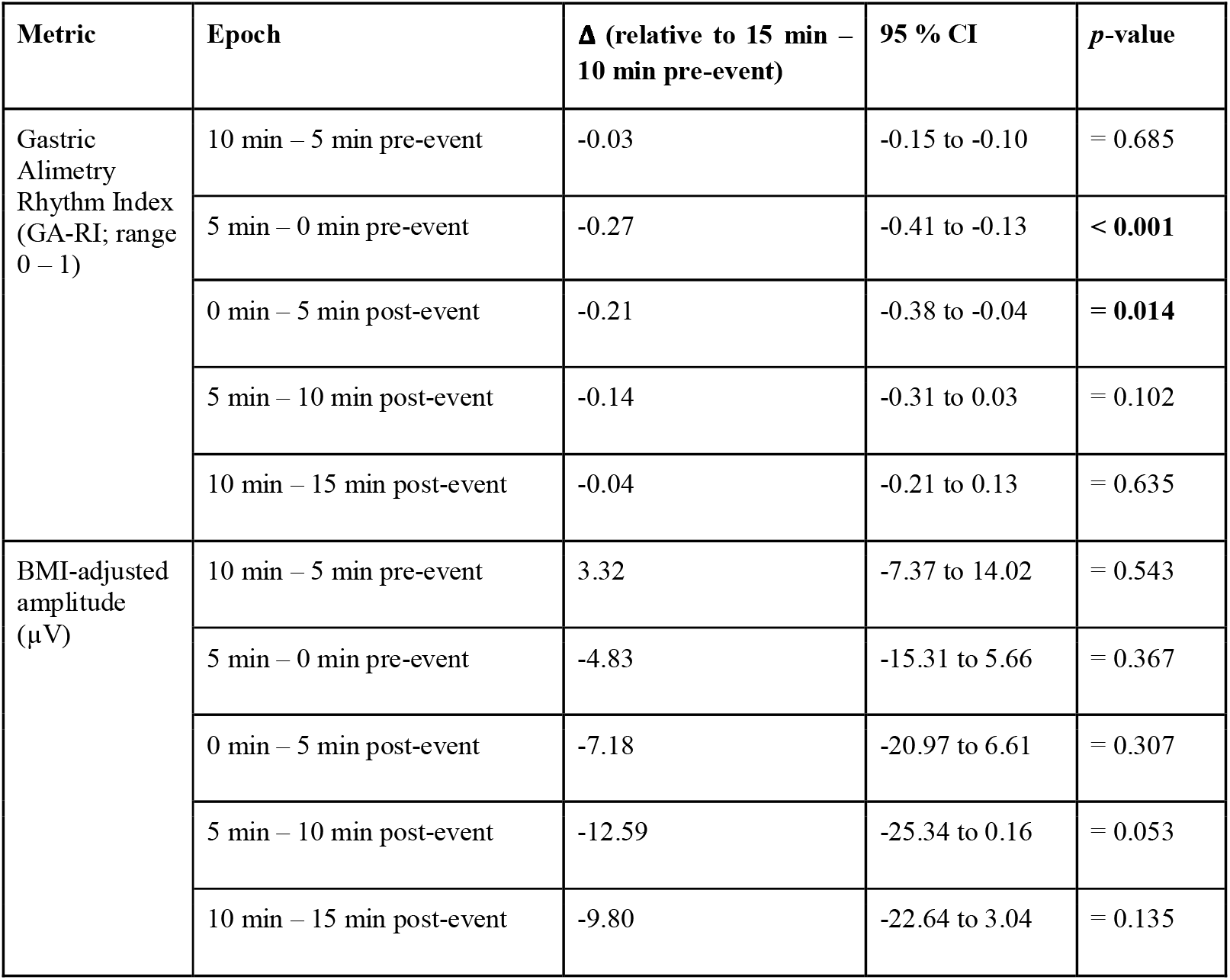
Mixed effects model of Gastric Alimetry Rhythm Index in pre- and post-vomit epochs.

In six cases, individuals with normal pre-vomit BMI-adjusted amplitudes dropped below the cut-off value (< 22 µV) after vomiting (**Fig. 3**). These individuals had BMI-adjusted amplitudes of 28.56 µV ± 5.45 µV during the reference period which dropped to a lowest value of 17.79 µV ± 3.76 µV after vomiting. In all of these tests, the BMI-adjusted amplitude recovered to above the cut-off value (22 µV) after mean 40 min (range: 5 min - 85 min).

**Fig. 3.**
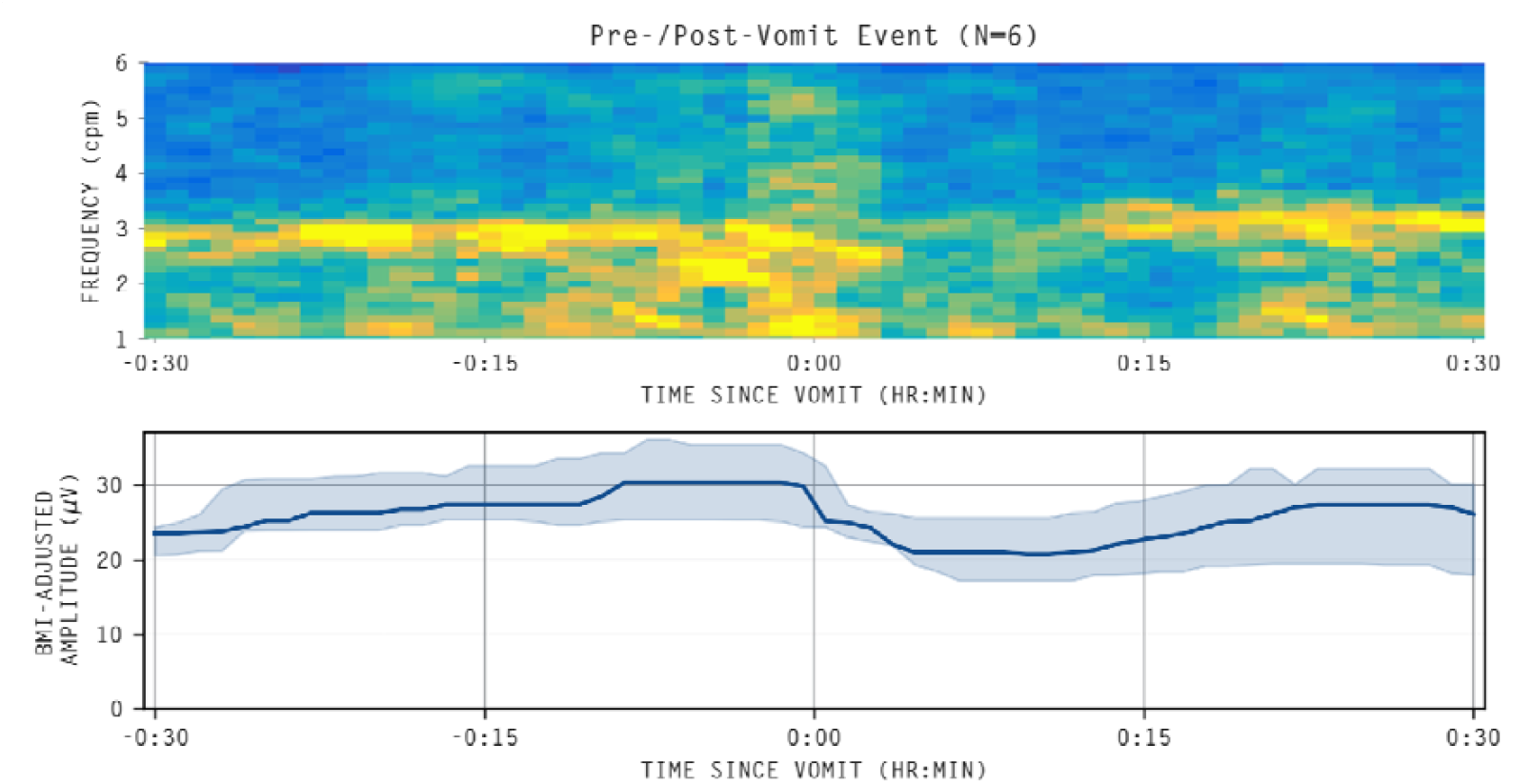
Average spectrogram across N=6 vomiting events (from 6 individuals) that were associated with a drop in BMI-adjusted amplitude below the reference range value (22 µV). In all of these tests, the BMI-adjusted amplitude recovered to above the cut-off value (22 µV) after mean 40 min (range: 5 min - 85 min)

### Symptomatic Relationship with Vomiting

A mixed effects model of symptoms over the vomiting period revealed that nausea, bloating, and excessive fullness each decrease following vomiting, in relation to the reference period (**Table 2**). Nausea was found to decrease in all post-vomiting epochs. Similarly, bloating decreased across all post-vomiting periods. Excessive fullness decreased in all post-vomiting epochs as well as during the 5 min immediately before vomiting. Vomiting was not associated with changes in heartburn, stomach burn, or upper gut pain measures.

**Table 2.**
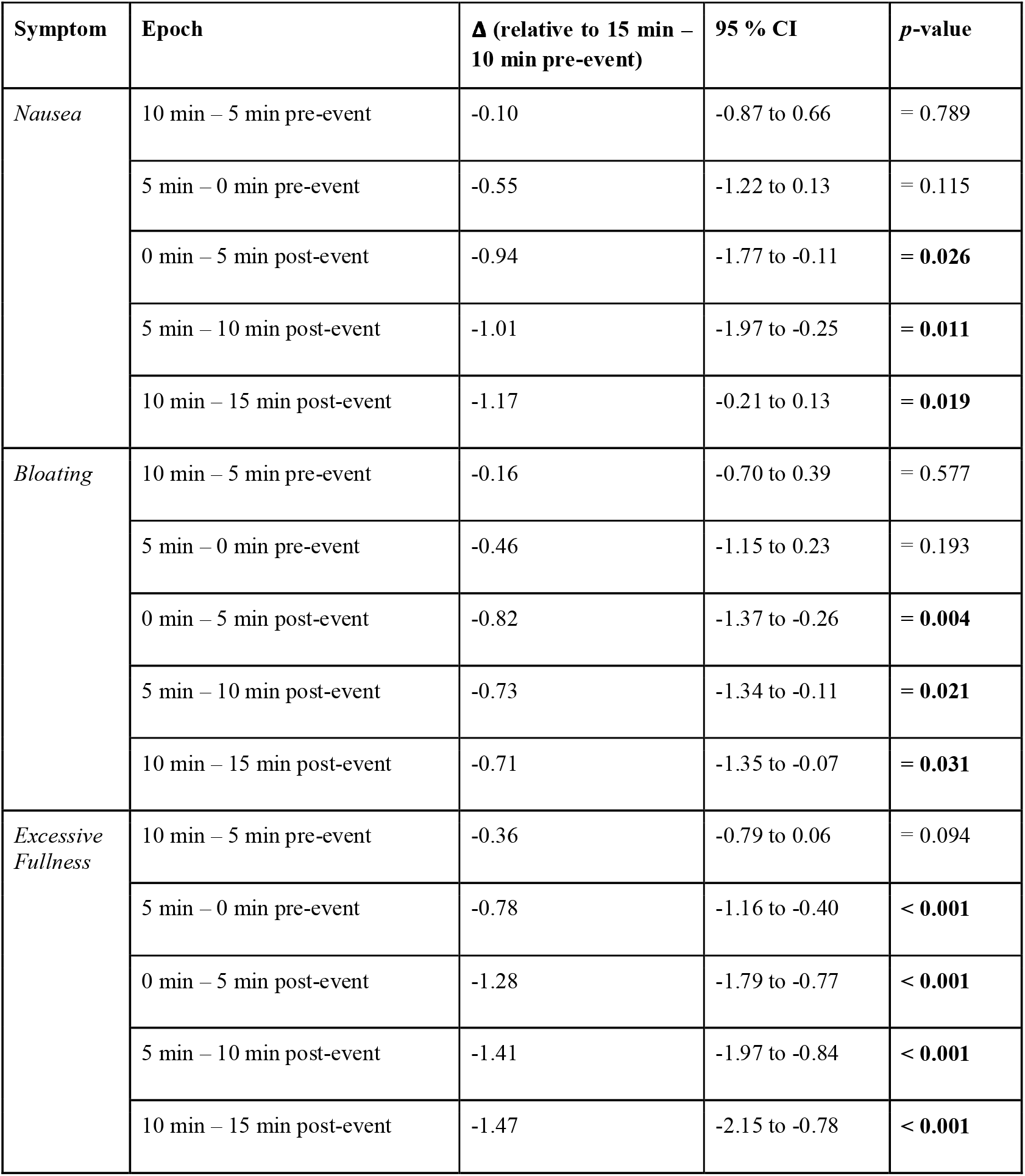
Mixed effects model of nausea, bloating, and excessive fullness symptoms in pre- and post-vomit epochs.

## Discussion

This study investigated the effect of vomiting on BSGM spectra and clinical test interpretation. In general, vomiting resulted in minimal visible changes to the BSGM spectrogram, with 90% of cases maintaining a qualitatively consistent bioelectrical profile throughout the event (*e*.*g*., **Fig. 1**). Vomiting was not associated with a typical phenotype or clinical presentation, though, as expected, vomiting was associated with symptomatic improvement for bloating, nausea, and excessive fullness.

Granular quantitative evaluation at high temporal resolution revealed that vomiting was associated with brief and transient decreases in GA-RI across an approximately 10 min period (5 min either side of the logged event) and occasional motion-related artifacts. Despite these localised effects, there was no distinct, consistent ‘vomiting signature’ on BSGM clinical reports, and importantly, vomiting did not typically alter a patient’s BSGM spectral phenotype, which (alongside symptomatic patterns) is principally used to diagnose pathophysiology.(Gharibans et al. 2022; O’Grady et al. 2023b; Varghese et al. 2023; Wang et al. 2024; Varghese et al. 2025b)

While transient vomiting effects did not affect the overall clinical or statistical presentation of any patient, there were 4 cases where vomiting soon after meal consumption was associated with a delayed meal response. These scenarios warrant careful consideration as they could confound clinical interpretation. Principally, a delayed meal response could indicate impairment to neurological signalling along the gut-brain axis, and/or autonomic inhibition of gastric slow wave activity.(Chan et al. 2021; Mercado-Perez and Beyder 2022) Alternatively, this association may have been incidental. Physiologically, vomiting reduces upper GI volume and calorie content, potentially attenuating the typical amplitude ramp seen in healthy digestion.(Huang et al. 2024) Other studies, however, have noted delayed meal responses as a pathophysiological feature without specific vomiting events in patients with gastroparesis-like symptoms(Chen et al. 1996) and chronic nausea/vomiting syndrome,(Gharibans et al. 2022) as well as in adolescents with chronic gastric symptoms.(Humphrey et al. 2025) Ultimately, such cases currently therefore require clinical interpretation and judgement; vomiting episodes are annotated (along with reflux and belching events) via the app-based patient interface and included in the final report, so that the clinician can determine their significance. Based on this study, our advice is that the presence and timing of logged vomiting events are taken into consideration during clinical evaluation of BSGM reports, and cautious interpretation of associated delayed meal responses is warranted until further data may become available.

It is well known that vomiting any portion of the radiolabelled meal during gastric emptying scintigraphy, a standard diagnostic test for gastroparesis, negatively impacts the validity of the test.(Abell et al. 2008) Since the integrity of the test relies on the passage of the standardized radiolabelled meal out of the stomach into the small intestine, vomiting even a portion of the meal out of the body reduces the reliability of the test, which may be terminated thereafter. By contrast, based on this study, it appears that the overall integrity of BSGM clinical tests are not significantly compromised by vomiting. Despite this, caution should be exercised when considering reference intervals for BSGM metrics (specifically, GA-RI and BMI-adjusted amplitude) when large-volume vomiting occurs, as well as tests with unsupervised vomiting events. This is because the reference intervals are based upon the fed stomach, which has certain stretch- and nutrient-mediated mechanisms influencing bioelectrical activity that is likely altered following vomiting, particularly events involving the evacuation of substantial proportions of the meal.(Levanon et al. 1998; Beyder et al. 2010) This can also be inferred by the observation that smaller meal ingestion is known to affect BMI-adjusted amplitude and GA-RI metrics.(Huang et al. 2024) Conversely, motion artefact noise at the time of vomiting is likely not an important clinical consideration, as significant noise was rare (6 % of cases), being short-lived (localised to a few minutes either side of the vomit episode), and successfully cancelled from metric calculations by the automated signal processing pipeline in all cases.(Gharibans et al. 2018; Calder et al. 2022) Additionally, though GA-RI dropped locally at the time of vomiting, there was no difference in PGF, BMI-adjusted amplitude, or GA-RI when data surrounding the vomiting event was omitted from the overall metric calculations, suggesting that the clinically-reported metrics were robust to vomit-related disturbances. It therefore stands to reason that the ability of BSGM to determine clinically-meaningful phenotypes and their underlying pathophysiology on a patient-by-patient level is unaffected by vomiting. This is perhaps best illustrated by **Fig. 2**, in which the clinical phenotype of each patient is still determinable, regardless of the effects of vomiting and the subsequent delayed meal response.

Previous studies have shown that consuming a calorie-reduced meal has not had a significant effect on BSGM test quality or clinical interpretation.(Huang et al. 2024) The present data further supports this conclusion, as vomiting a portion of the meal did not significantly affect the usability and application of the BSGM test. A smaller meal may therefore be a suitable alternative intervention for patients with a history of frequent post-prandial vomiting, though careful consideration of the metrics’ reference ranges is warranted under these conditions.

Various literature, particularly from canine studies, has highlighted the presence of gastric slow wave dysrhythmias at the time of vomiting,(Lang et al. 1986; Ueno and Chen 2004; Yu et al. 2009) and it has been hypothesised that these transient electrical patterns may play a functional role in the physiology of vomiting, for example by temporarily disrupting normal gastric motility patterns. This hypothesis is supported by the present data, which found a statistically significant decrease in GA-RI, localised to a 10 min period surrounding the logged vomiting event. These physiological changes, which suppress normal motility, are likely to be autonomically-mediated and are considered to help to prepare the stomach to receive and accommodate retropulsed small intestine contents, prior to their forceful oral evacuation.(Lang 2023)Importantly, the temporal scale with which this event occurs is too small to be reflected in the overall clinical reports automatically generated following a Gastric Alimetry test, and this dysrhythmia is relatively suppressed in standard clinical data presentations due to smoothing and averaging effects of the test metrics. However, these physiological changes also more generally highlight that gastric slow wave dysrhythmias correlate with vomiting in humans. Importantly, the very brief and transient nature of these dysrhythmias also means vomiting effects will not confound the diagnosis of more prolonged or continuously dysrhythmic states that have been linked to gastric motility disorders such as interstitial cell of Cajal pathologies.(Gharibans et al. 2022; Varghese et al. 2025b)

A limitation of this study was that it evaluated a de-identified BSGM database available for clinical research, without linkage to medical histories, in order to focus on the electrophysiology of human vomiting. As such, underlying diagnoses and aetiologies attributed to vomiting disorders are not described here. However, no patients were on medications affecting gastric motility at the time of testing, in accordance with standard test protocols.(O’Grady et al. 2023a) Further, an assumption was made that of the 49 logged vomiting events that were analysed, none were false negatives. This reflects a limitation in the retrospective nature of the study, whereby vomiting events could not be confirmed for validity against study notes or medical history. Furthermore, we were unable to ascertain the volume of vomitus ejected for any given event. However, the App includes a validated pictogram and text interface that has previously been validated as being specific as a patient reported outcome tool for vomiting events.(Sebaratnam et al. 2022) Further vomit-specific information, such as volume and contents, would have been desirable to strengthen the present analyses. In the future, an examination of repeat tests with and without vomiting events may offer additive insights, however, this would likely require a vomiting induction protocol which poses ethical and recruitment challenges.

In conclusion, vomiting during BSGM testing was associated with transiently decreased amplitude and rhythmic stability within 5 min pre- and post-event. Subsequently, these temporary changes normalised, and there was minimal impact on the overall 4.5 hr test summary metrics. While additional noise and atypical responses around the emesis may require consideration during clinical interpretation, this study finds that the overall impact of vomiting on BSGM test electrophysiological metrics is small. BSGM therefore overcomes a significant limitation of gastric emptying tests and is a suitable diagnostic tool for characterising conditions with persistent vomiting.

## Data Availability

All data produced in the present study are available upon reasonable request to the authors.

